# Spatially and Temporally Precise Microbiome Profiling in the Small Intestine using the SIMBA Capsule with X-ray tracking

**DOI:** 10.1101/2024.04.02.24305212

**Authors:** Gang Wang, Sharanya Menon, Lynn Wilsack, Renata Rehak, Lawrence Lou, Christian Turbide, Jeremie Auger, Annie Tremblay, Olivier Mathieu, Sylvie Binda, Thomas A Tompkins, Sabina Bruehlmann, Christopher N Andrews

**Affiliations:** Nimble Science Ltd., Calgary, Alberta, Canada; University of Calgary, Cumming School of Medicine, Calgary, Alberta, Canada; Rosell Institute for Microbiome and Probiotics, Lallemand Health Solutions, 6100 Royalmount avenue, Montreal, Quebec, Canada; Lallemand Health Solutions, 19 Rue des Briquetiers, BP 59, 31702 Blagnac, France; Lallemand Bio-Ingredients, 6100 Royalmount avenue, Montreal, Quebec, Canada

**Keywords:** gut microbiome, small intestine, probiotics, ingestible sampling capsule

## Abstract

Few minimally invasive options for sampling the small intestinal (SI) luminal fluid exist to study the SI microbiota in health and disease. To address the lack of tools and methods to study GI regions that are difficult to access, Nimble Science developed a fully autonomous and passive sampling method, the Small Intestine MicroBiome Aspiration (SIMBA^TM^) capsule, for convenient, high-quality, and reliable sampling to study the diet-microbiota interactions in the SI. The sealing efficacy and microbial DNA preservation capacity of the SIMBA capsules was first validated through *in vitro* simulation assays. Then, a clinical study was conducted with 20 healthy participants to validate the *in vivo* use of SIMBA capsules to reliably capture samples for SI microbiome analysis before and after an intervention (NCT04489329). Briefly, participants ingested the capsules at baseline and 7 days later, with a probiotic capsule containing a blend of *L. rhamnosus* R0011 and *B. longum* R0175. Following baseline SIMBA capsule ingestion, multiple low-dosage x-ray scans were performed to track the sampling location. Fecal samples corresponding with the baseline and intervention capsule were analyzed for comparison. The SIMBA capsules’ performance *in vitro* demonstrated the potential for contamination-free sampling with preservation of the microbial communities. Within the clinical study, the capsules performed safely and reliably for collection of SI content. X-ray tracking confirmed that 97.2% of the capsules completed sample collection in the SI regions before reaching the colon. Importantly, our data showed that the capsules sampled in the right area of the intestines and that baseline SIMBA microbiome profile is significantly different from fecal microbiome profile. SIMBA successfully detected a concurrent probiotic intervention in the small intestine, which was not detectable using stool samples. The high accuracy of sampling location and sealing efficacy of the SIMBA capsules makes them potentially useful research tools in clinical trials for studying diet-microbiota interactions in health and disease, and perhaps eventually for the clinical diagnosis of GI tract conditions affecting the SI such as SIBO.

## Introduction

The distribution of microorganisms varies along the length of the gastrointestinal tract, with specific niches influenced primarily by pH, oxygen levels, antimicrobial peptides, and bile acid and nutrient gradients.(1) The bacterial density increases from 10^1^ – 10^3^ cells/gram in the stomach and duodenum, to 10^4^ – 10^8^ in the jejunum and ileum, reaching 10^11^ – 10^12^ in the colon.(2) Furthermore, the acidic pH and oxygen-rich environment of the proximal small intestine favors microbial colonization with acid- and oxygen-tolerant bacteria (e.g. *Lactobacillus*, *Streptococcus*, *Veillonella*), whereas in the colon, oxygen-poor conditions and slower transit results in fermentation of complex polysaccharides, resulting in greater taxonomic diversity and dominance of saccharolytic anaerobic *Bacteroidales* and *Clostridiales*.(3)

Perturbation of microbial composition of the normal gut microbiome (dysbiosis) is associated with a number of gastrointestinal disorders and disease states. In the small intestine, dysbiosis is associated mainly with small intestinal bacterial overgrowth (SIBO), a condition often intertwined into the symptomatology of other functional bowel diseases such as irritable bowel syndrome, inflammatory bowel diseases, or short bowel syndrome.(4) SIBO incidence can be increased in the presence of dysfunctions of SI protective mechanisms (e.g. lower antimicrobial gastric and biliary secretions, reduced anterograde peristalsis preventing microbial adherence, loss of the ileocecal valve inhibiting retrograde translocation of colonic bacteria, reduced mucin production by mucosal epithelial cells trapping bacteria, reduced cellular/humoral immunity and anti-bacterial peptides), particularly in the setting of systemic illness, altered anatomy (e.g. surgery), or medications.(5, 6)

The location and length of the small intestine makes direct microbial sampling challenging. While an endoscopically attained duodenal aspirate with a bacterial count > 10^5^ CFU/ml has historically been considered the gold standard for diagnosing SIBO, criticisms include the relative invasiveness, patient risk, cost, as well as technical factors including the lack of standard approach, difficulty in preserving anaerobic environment, concern of microbial contamination in the proximal gut, and the chance of missing bacterial overgrowth in the mid-distal small bowel.(7) The introduction of 16S RNA gene amplification/metagenomic analysis does not rely on the culturability of bacteria and can allow for the profiling and characterization of the microbiome in the sample.(8) A prospective study of 15 symptomatic patients who underwent jejunal aspirate with subsequent 16S rRNA gene sequencing and metagenomic analysis, demonstrated differing bacterial compositions between the jejunum, colon, and oropharynx, which confirmed the findings of previous autopsy studies.(9, 10)

Breath testing, which involves ingestion of a prespecified carbohydrate substrate (generally glucose or lactulose) with subsequent quantification of exhaled gasses (generally hydrogen and methane, which are hydrolyzed by colonic bacteria) at regular intervals, has been proposed as an indirect diagnostic tool for SIBO. The presence of a premature rise in exhaled hydrogen is suggestive of colonic bacteria moving proximally to the distal small intestine.(11) However, this too has criticisms; glucose is absorbed in the proximal small bowel, resulting in decreased sensitivity, and the peak associated with lactulose has been demonstrated to be a marker of oro-cecal transit.(11–13) Furthermore, up to 10% of patients may not produce hydrogen after carbohydrate ingestion, due to colonization with methane-producing organisms.(14) Ultimately, the evaluation of the small intestine for research and clinical purpose, including the diagnosis of SIBO, is fraught by the lack of a validated gold standard.

These challenges are also confounding research on diet-microbiome interactions associated with other conditions related to the small intestine, such as IBD, obesity, metabolic disease and cancer.(5–7, 9) Further, the evaluation of the restorative impact of therapeutic interventions, including probiotics is hampered by the lack of accessibility to the delivery site.(15)There is a critical need for minimally invasive devices that allow direct high-quality sampling in the small intestine. Over the past few years, a few capsule-based sampling systems were introduced to target the small intestine using passive sampling technologies.(16–18) While an early study in humans has demonstrated luminal fluid collection with multi-omic profiles differing from stool,(16) these technologies have overlooked the significance of effective sealing in the SI and preserving sample quality. Further, this is the first report of spatial and temporal accuracy, including use in humans with and without intervention. In this study, we developed a pH-based autonomous and passive Small Intestine MicroBiome Aspiration (SIMBA) capsule. The SIMBA capsules have unique features of large sampling ports for reliable sampling volume, strong sample sealing performance and embedded microbial DNA preserving agents to ensure sample quality during the capsule transit time and capsule return process. The results of *in vitro* simulation assays and clinical study presented herein confirm that SIMBA is a well-tolerated, minimally invasive capsule that passively captures, seals, and preserves small intestine luminal fluid, providing samples that are suitable for downstream microbiome analysis.

## Methods

### SIMBA Capsules

The capsule has overall dimensions of 25.4 mm (L) × 8 mm (D), the size of a 00EL capsule. The functional device is contained within a pH sensitive small intestine targeting outer shell. The outer shell is designed to disintegrate at a nearly neutral pH, which is similar to the pH of the proximal small intestine region. The capsule comprises a main body which includes a sampling chamber with sufficient volume (∼105 µL) to capture a representative sample of intestinal fluid. The intestinal fluid enters the chamber through four evenly spaced ports which are accessed by the fluid only upon disintegration of the outer shell (Figure 1).

**Figure 1:**
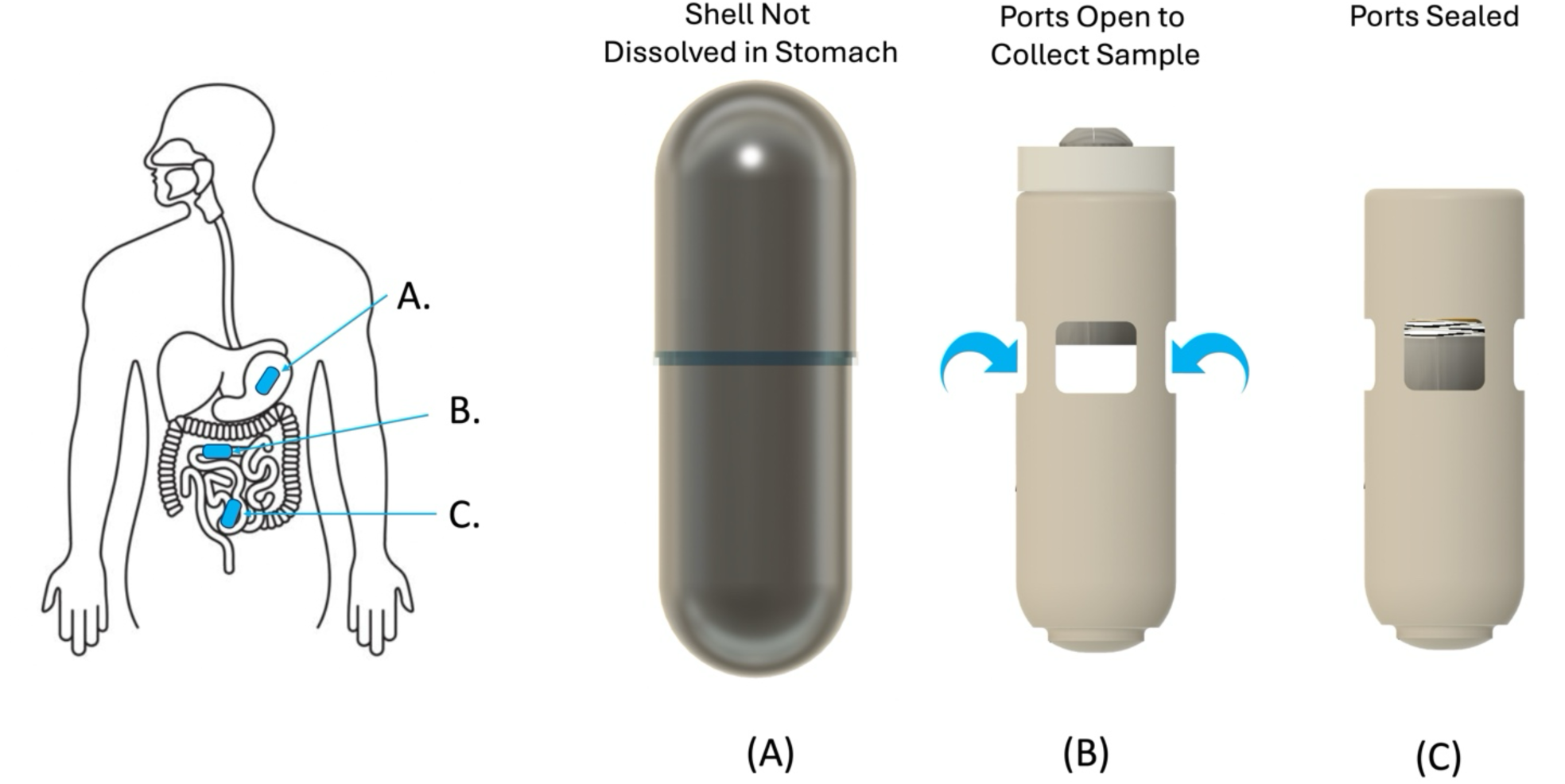
Configuration of the SIMBA capsule as it travels along the gastrointestinal tract. The outer shell of the capsule remains intact in the stomach (A), and dissolves as it reaches the proximal small intestine, where it opens and collects a sample of the surrounding fluid (B). The capsule then closes and remains sealed throughout the GI tract until expulsion and sample retrieval in the lab (C).

The ports are radially facing towards the mucosal layer of the small intestine to collect luminal samples near the mucosa. The ports are sufficiently wide to allow easy inflow of the liquid sample. Hydrophilic fibers are placed in the chamber to wick in and retain the liquid sample. On the top of the sampling chamber, a piston is held in the open position by a latch until it is exposed to small intestine fluid and dissolved in a time-controlled manner after the outer shell dissolution. Then, the sampling chamber is closed and sealed with a compressive spring forced by the piston on its top end.

Importantly, a preserving agent is embedded within the sampling chamber to maintain the integrity of the collected sample during the capsule transit time in the gut and the capsule return process. The sampling chamber is closed on the lower end by a cap. After retrieval of the device, the cap is easily removed to access the collected sample for downstream processing and analysis.

### *In vitro* sealing performance and preserving agent efficacy

To assess the sealing performance, 37 SIMBA capsules without outer shells were submerged in sterile PBS for 4 hours, automatically triggering the sampling and sealing mechanism in the presence of an aqueous solution. 34 capsules were then transferred to a healthy donor fecal slurry spiked with *L. rhamnosus* R0011 (∼10^9^ CFU/mL). Out of these 34 capsules, 3 were manually unsealed as a positive contamination control. The 3 remaining capsules were not exposed to the fecal slurry. All the control capsules and the treatment SIMBA capsules, while being submerged in the fecal slurry, were anaerobically incubated at 37 °C for 72 hours (to simulate the gut transit environment), then at 4 °C for 72 hours (to simulate a cold shipping condition). Then, the capsules were opened, and samples were recovered by pipetting. Contamination was assessed using a strain-specific SYBR Green qPCR assay targeting R0011.

Another set of 20 capsules were subjected to the same sealing procedure in PBS followed by a 24 h incubation in R0011-spiked healthy donor fecal slurry. While these capsules remained in the slurry, 8 capsules were frozen at -20 °C for 24 h or 168 h, and 2 capsules were incubated at 4 °C for the same time points (Table 1). After thawing, capsules were removed from the slurry and then samples were retrieved from the capsules by pipetting and contamination was assessed with a strain-specific SYBR Green qPCR assay targeting R0011.(19)

**Table 1:** *In vitro* sealing performance testing.

| Experimental condition | R0011<br>positives/total<br>number of capsules | Sealing<br>Efficacy |
| --- | --- | --- |
| Exposed to fecal slurry, sealed, 72h@37 °C + 72h@4 °C | 2/31* | 93.5% |
| Exposed to fecal slurry, unsealed, 72h@37 °C + 72h@4 °C | 3/3 | n/a |
| Not exposed to fecal slurry, sealed, 72h@37 °C + 72h@4 °C | 0/3 | n/a |
| Exposed to fecal slurry, sealed, 72h@ - 20 °C | 0/8 | 100% |
| Exposed to fecal slurry, sealed, 72h@ 4 °C | 0/2 | 100% |
| Exposed to fecal slurry, sealed, 168h@ -20 °C | 0/8 | 100% |
| Exposed to fecal slurry, sealed, 168h@ 4 °C | 0/2 | 100% |
| All test conditions | 2/51* | 96.1% |
| *The 2 positives are due to mishandling of the capsules before/during retrieval of the cargo and not contamination through the sealing mechanism. |  |  |

For preserving agent efficacy testing, fresh SI endoscopic aspirates from 3 patients collected on the same day from the Intestinal Inflammation Tissue Bank (IITB), University of Calgary were pooled, homogenized and inoculated into 30 capsules containing the embedded preserving agent and 4 capsules without the preserving agent. The filled capsules were then sealed and incubated at 37 °C for 144 hours (6 days) under anaerobic conditions before sample retrieval for DNA extraction.

### DNA extraction and quantification

Once the capsules were received by the lab, the samples were removed from the capsules using a sterile pipette to avoid cross-contamination by the fecal matters attached to the outer surface of the capsule body. The samples from SIMBA and their associated control samples collected at various timepoints were stored at -20 °C for up to 7 days until the scheduled DNA extraction. DNA Extraction of SIMBA and fecal samples was performed with the Qiagen QIAamp PowerFecal Pro DNA Kit, following the manufacturer’s protocol with modifications for the SIMBA samples. The Qubit dsDNA HS Assay Kit was used with the Qubit® 2.0 Fluorometer to measure the SIMBA aspirate and fecal sample DNA concentration.

### 16S library preparation and sequencing for in-vitro preserving agent efficacy testing

For the preserving agent efficacy testing, the 16S rRNA gene V4 variable region was amplified using PCR primers with internal barcodes (primers: F: AATGATACGGCGACCACCGAGATCTACAC- barcode-TATGGTAATTGTGTGCCAGCMGCCGCGGTAA, R: CAAGCAGAAGACGGCATACGAGAT-barcode- AGTCAGTCAGCCGGACTACHVGGGTWTCTAAT) in a 35 cycle PCR using the KAPA HiFi HotStart master mix (Roche Sequencing). The conditions for the thermocycler were as follows: 98 °C for 2 minutes, followed by 35 cycles of 98 °C for 30 seconds, 55 °C for 30 seconds and 72 °C for 20 seconds, after which a final elongation step at 72 °C for 7 minutes. Amplified PCR products were checked in a 1 % agarose gel. The PCR products were then purified using NucleoMag NGS Clean-up and Size Select (Macherey-Nagel) and concentrations normalized using SequalPrep Normalization Plate (Invitrogen). Amplicons were pooled and concentration and quality were determined using the Qubit HS DNA kit (Invitrogen) and the Tapestation D1000 assay (Agilent), respectively. Amplicon sequencing was done on a MiSeq Benchtop DNA sequencer (Illumina) using a V2-500 cycle kit (Illumina Inc). The pooled library was then denatured and prepared for loading on an Illumina MiSeq cartridge with a 5% PhiX Control.

### Clinical study design, participants, and probiotic intervention

A clinical study was conducted to validate the ability of the SIMBA capsule device to collect samples from the small intestine for microbiome analysis in adults and to detect microbiome changes due to dietary interventions from simultaneous ingestion of a probiotic. The study was conducted under the guidance and approval of the University Conjoint Ethics Review Board (REB 20-1211) and Investigational Testing Authorization from Health Canada (ITA 318739). The clinical trial protocol was prospectively registered on Clinicaltrials.gov (NCT04489329).

All potential participants in the study had a screening visit or telephone call to review inclusion and exclusion criteria, review the consent form (or be sent an electronic copy of the consent form for review), and book the subsequent testing. No reimbursement other than parking costs were made to the participants. The main exclusion criteria include a history of a small intestine obstruction or symptoms of an intermittent small intestine obstruction, known history of IBD, swallowing disorders, and intestinal strictures, prior gastrointestinal surgery which had altered the gastrointestinal anatomy, use of any medications that could substantially alter gastrointestinal motor functions in the previous 7 days, use of antibiotics, prebiotics, herbal supplements, or probiotics for 2 weeks before the first visit or during the study.

On the morning of the first day (termed the Baseline day), fasted participants visited the X-ray facility and then ingested two SIMBA capsules simultaneously. After ingesting the capsules, low-dose (70-80kVp), multiple X-ray imaging was used to confirm the timing and location of the capsules. The protocol specifies a scanning interval of approximately 30 minutes, and each volunteer is not permitted to undergo more than 12 scans on the same day. Once capsules were seen finished sampling on an X-ray image, participants were provided with capsule and stool collection kits and discharged. Participants were then allowed to leave the clinic and resume normal activities.

Participants were asked to resume normal eating 4 hours after the capsule ingestion and maintain a stable diet until the second visit. Participants were asked to monitor their stool for the passing of the capsule, and upon excretion they were asked to collect the capsules and a stool sample from the same bowel movement using the provided retrieval kit and return it promptly in an ice box for analysis.

After at least 5 days, but no more than 21 days following Baseline day, participants returned for a second visit (termed Intervention day) at the Investigators office or a clinic. Participants were required to fast overnight in advance of this visit. In the morning, they first ingested the probiotic capsule (40 billion CFUs, containing a blend of *L. rhamnosus* R0011 (71%) and *B. longum* R0175 (29%), provided by Lallemand Health Solutions) under instruction and then immediately ingested two SIMBA capsules under supervision. Participants did not undergo X-ray monitoring on their second visit. The participants were allowed to resume normal activities and were allowed to eat 4 hours after the ingestion of the SIMBA capsules. Participants were provided with stool and capsule collection kits and were instructed again on procedures for capsule return. Upon excretion of the capsule, patients collected the capsules and a stool sample from the same bowel movement and returned it promptly for analysis.

### 16S library preparation and sequencing for Clinical Samples

The 16S targeted amplicon sequencing library was prepared by amplifying 10 µL of each capsule gDNA extracts (or 25 ng for the fecal extracts) with 1X KAPA HiFi HotStart ReadyMix (Roche, cat # KK2802) and 200 nM universal 16S primers (forward 5’-CCTACGGGNGGCWGCAG-3’ and reverse 5’-GACTACHVGGGTATCTAATCC-3’) targeting V3-V4 regions in a 25 µL reaction volume (20). PCR products were visualized on a 2% agarose precast E-Gel stained with SYBR Safe dye (Invitrogen cat # G72080). Amplicons were purified with Agencourt AMPure beads (Beckman Coulter, cat # A63881) following Illumina 16S Metagenomic sequencing library preparation’s protocol. A second round of amplification using 5 µL of the purified amplicon PCR reaction as template, 2.5 µL each of Nextera XT V2 primers sets A and D (Illumina, cat # FC-131-2001 and FC- 131-2004) and 1X KAPA HiFi ReadyMix was performed in 25 µL reactions with the same cycling conditions as the Amplicon PCR except that only 8 cycles were used. PCR reactions were again purified with AMPure beads before individual fluorescent quantification by Quant-iT PicoGreen dsDNA assay (Life Technologies, cat # P7589). Volumes corresponding to 100 ng of each purified Index PCR reaction were pooled using an EpMotion 5075 liquid handling robot (Eppendorf) and this pool was quantified with QuBit Broad Range assay (ThermoScientific, cat # Q32853) following manufacturer’s instructions. This pool was also quality controlled for the presence of the desired amplicon (size obtained 630 bp) and the absence of secondary amplification by running a High Sensitivity D1000 TapeStation assay (Agilent, cat # 5067-5584/5585). Library was denatured with 0.2 N NaOH and loaded at 8 pM with 5 % PhiX (Illumina, cat # FC-110-3001) on an Illumina MiSeq instrument using MiSeq V3 Reagent Kit (Illumina, cat # MS-102-3003) for 2x 301 cycles.

### Microbiome analysis

The demultiplexed fastq sequences were imported into QIIME2 (Quantitative Insight Into Microbial Ecology–2) as artefacts and inspected for overall quality (visual inspection of the q-Scores per base plots). The reads were determined to be very high quality on the 40nt->280nt for the forward and 40nt->260nt for the reverse reads. These parameters were used to denoise the paired reads using the Dada2 denoiser (as a QIIME2 encapsulated version). Hence, the reads were clustered into amplicon sequence variants (ASVs). The feature classifier was used to attribute the ASVs to the closest known taxa using QIIME2’s taxonomic classification module (linking ASV sequences to known bacterial groups). The taxonomy file was trained on a 99% clustered Silva_138 taxonomic database (V3-V4 subregion of the 16S). The ASV tables were exported as Level-6 (Genus Level) relative abundance tables for downstream analysis.

The ‘core-metrics’ module from QIIME2 was also used to generate the Alpha Diversity measures and the PCoA distance matrices. The alpha diversity algorithms include Pielou (Evenness), Faith (Phylogenetic Distance) and Shannon Entropy. For the PCoA, the Weighted UniFrac algorithm was used. The PCoA was viewed interactively with the Emperor module through the QIIME 2 viewing server (https://view.qiime2.org/) and a collection of images was captured for later reporting. The PCoA and diversity figures were calculated on ASV tables and rooted tree (phylogenetic relation between observed ASV sequences).

To determine group differences between the treatment groups, QIIME2’s encapsulated Machine Learning Sample Classifier was used with the ExtraTreesClassifier algorithm. In order to assess the presence or absence of differences between treatment groups (or Capsules V. Stool samples), the algorithm trains on 2/3 of the sample’s taxonomic tables at genus level (Training Set) and then test its predictive power on the remaining 1/3 of the samples from each group (Testing Set). If the algorithm is able to tell the samples from each other for the Label of interest (the variable used to make the groups, ex. Stool V. Capsule samples), then the groups are determined to be different. The accuracy results of the Test Set results are presented as confusion matrices, with the main classification indicator being the Final Accuracy.

## Results

### SIMBA can effectively seal and protect collected samples

The goal of our in vitro validation experiments was to assess the possibility of contamination occurring after completion of the sample collection and sealing of the SIMBA capsule, simulating the conditions of capsule transit through the GI tract and potential shipping and storage conditions. For the first experimental design, there was no contamination in 29 out of the 31 test capsules as shown by the absence of detection of *L. rhamnosus* R0011 in the capsules’ cargo. The 2 samples with positive detection of R0011 are the result of one obstructed capsule preventing it from completely closing (which precludes from assessing sealing efficacy) and one capsule that incidentally touched the biosafety cabinet during the sample removal process, the cargo coming into contact with the outside of the capsule. Therefore, the detection of R0011 in these 2 samples was not due to failure of the seal and were excluded from the sealing efficacy evaluation (Table 1). Importantly, no target bacteria were detected in the negative controls, and R0011 was detected between 10^4^ and 10^5^ in the contamination positive controls (i.e., manually unsealed before immersion in the contaminating slurry).

In the second set of sealing efficacy testing (Table 1), all 16 frozen samples were negative for R0011 after thawing, regardless of the time spent at -20 °C. This temperature was tested to ensure that the spring-based mechanism and sealing capacity of the capsules were able to withstand freezing stress causing an expansion of the cargo. The 4 samples kept at 4 °C were also negative for R0011. Overall, the sealing efficacy of the SIMBA capsule was conservatively evaluated at 2 positives/51 tested capsules (96.1 % efficacy) (Table 1).

Most of the 16S microbiome sequencing profiles of the endoscopic aspirate samples stored in the SIMBA capsules with the preserving agent (Group P, N=30) are similar to the T0 control (Figure 2A), while the 4 samples without the preserving agent (Group NP, N=4) are characterized by a notable dominance of *Pasteurellaceae*. The absence of preservative allows for the overgrowth of some species, which changes the relative abundance in the sample. Among samples with the preserving agent, 5/30 capsules showed a variable amount of staphylococcus contamination after 6 days at 37 °C. After removal of *Staphylococci* by bioinformatics filtering, the 5 samples displayed a 16S profile similar to the other samples and T0 (Figure 2B).

**Figure 2:**
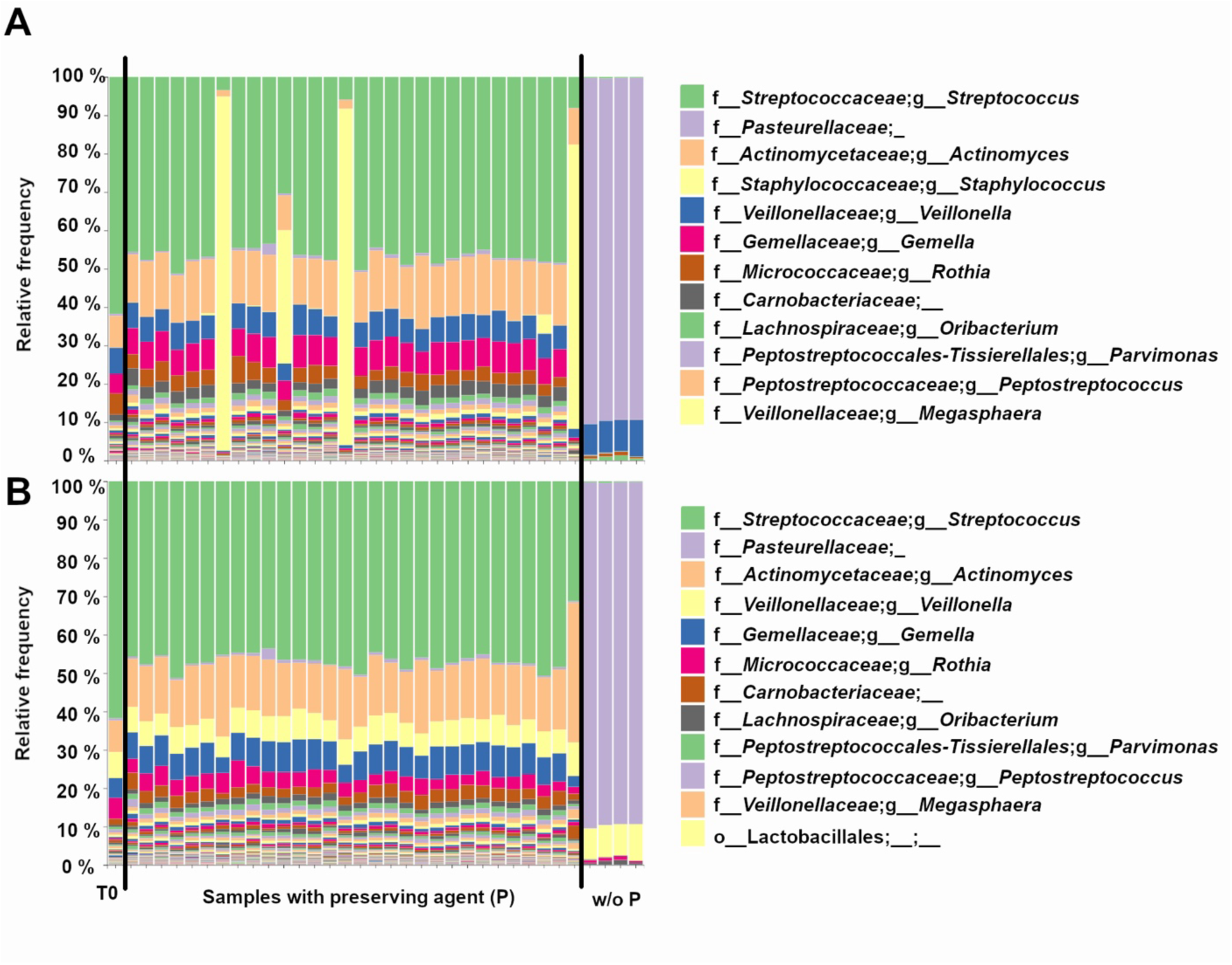
Bar plots showing the genus-level 16S microbiome sequencing profiles of endoscopic aspirate samples that were (A) inoculated in the SIMBA capsules for the T0 control, the samples with preserving agent (N=30) and without preserving agent (w/o P) (N=4). (B) same as in (A) but with bioinformatics filtering to remove the *Staphylococci*.

### SIMBA Performs Safely and Reliably for Collection of Small Intestine Luminal Fluid *in vivo*

The clinical study results are summarized in Table 2. This single arm study enrolled 20 healthy volunteers (mean age (±SD): 38 ± 11, range 18-58 years; mean BMI 25.6 ± 4.7; 12 females and 8 males) recruited at the Foothills Hospital in Calgary Alberta. The SIMBA capsules were well tolerated by participants (N = 20), with no adverse events or capsule retention, and only 5 out of the 40 incidences (2 capsule retrievals by each of the 20 participants) where participants judged the first-time retrieval of the capsules from stool in the baseline round was “difficult”, 18/40 “Neutral”, 13/40 “easy”, 4/40 “very easy”, and 1/40 “Lost”. In the probiotic round, the difficulty of capsule retrieval was reported as follows: 5/40 “difficult”, 11/40 “Neutral”, 18/40 “easy”, 3/40 “very easy”, and 1/40 “Lost”. Overall, 78/80 (97.5%) capsules (39 baseline + 39 probiotic intervention) were retrieved by participants, after a median number of 2 stools (range 1-7) and with total median transit time (ingestion to expulsion) of 30 hours (IQR 23-48). The two missing capsules were established as lost in feces in two participants through follow-up x-ray scans confirming capsule clearance.

**Table 2:** Summary of the clinical study results.

| Tasks | # Capsules | # Ingestion Events | # Participants |
| --- | --- | --- | --- |
| **User-level success rate** |  |  |  |
| Swallowing w/o Difficulty | 80 / 80 | 40 / 40 | 20 / 20 |
| Capsule Ingestion | 80 / 80 | 40 / 40 | 20 / 20 |
| Independent Capsule Retrieval | 78 / 80 | 40 / 40 | 20 / 20 |
| **Capsule performance** |  |  |  |
| Capsule Transit |  |  |  |
| within 72 hrs | 76 / 78 | 39 / 40 | 19 / 20 |
| within 96 hrs | 78 / 78 | 40 / 40 | 20 / 20 |
| Sample Processing Without Contamination | 78 / 80 | 40 / 40 | 20 / 20 |
| Small Bowel Targeting Accuracy (Baseline only) | 35 / 36* | 19 / 20 | 19 / 20 |
| * 4 indeterminate due to X-ray schedule. |  |  |  |

Baseline capsules were monitored by X-ray (2 capsules per participant) following ingestion to determine the sampling time and region. Figure 3A and 3B show examples of the full abdominal X-ray images taken from one participant at two different timepoints. Figure 3C highlights the region of interest (ROI) in which the two capsules were still open (sampling); Figure 3D shows the ROI in which the two capsules were closed (sampling completed). A radiopaque marker was attached on one of the two capsules ingested together in order to distinguish the two capsules in the X-ray images. In total, 35/40 (87.5%) capsules were confirmed as having completed sample collection in the targeted region of the small intestine. The only failed case was a capsule that was seen completing collection in the stomach before reaching the small intestine. Four capsules were classified as indeterminate due to the X-ray scanning frequency or the overall duration of the X-ray scanning period. Of these four capsules, two were still in the stomach at the end of the X-ray schedule, although sampling collection was not completed. Two other capsules were last seen open in the small intestine in one X-ray scan and were first seen closed on the next scan but had already reached the colon. If these four cases were excluded from the sampling location analysis, 35/36 (97.2 %) capsules were observed collecting samples in the small intestine before reaching the colon, with 1/36 (2.8 %) completing its collection while still in the stomach. Overall, 38/40 capsules completed sample collection within 210 minutes following ingestion (IQR 150-180 minutes).

**Figure 3:**
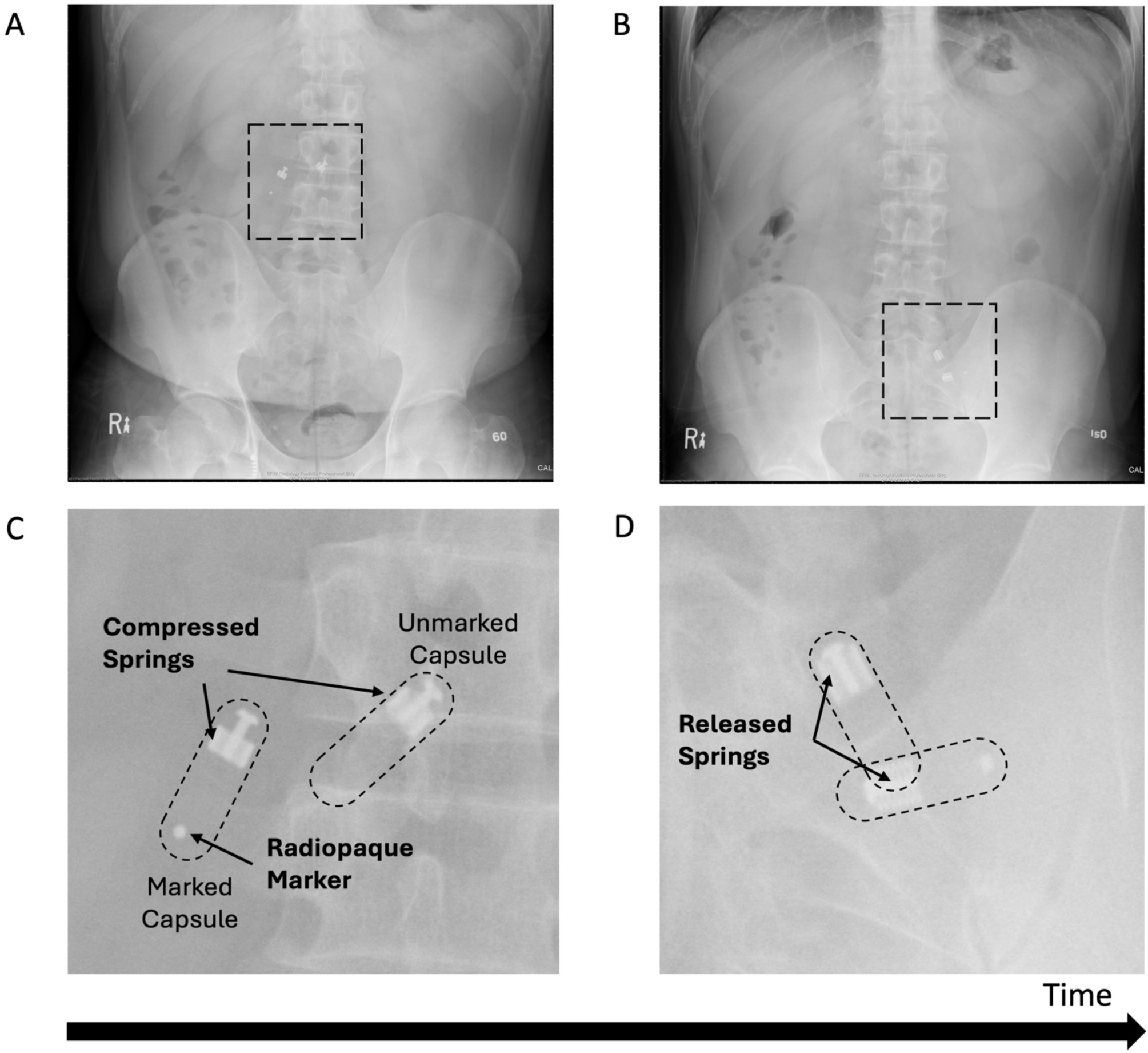
*In situ* X-ray images. Capsules are in the compressed configuration on the full X-ray image (A) and corresponding ROI (C) while sampling, and on the full X-ray image (B) and corresponding ROI (D) in the released, sealed configuration after sampling is completed. Representative images are from 1 participant.

The mean sample weight per capsule was 89 ± 27 mg (range 15-130 mg). In total, 87.2% (68/78) of the SIMBA capsules collected more than 20 mg of sample, which is the threshold weight we set to assess the sample collection efficacy. However, the sample collection rate increased to 97.5% (39/40) by ingestion under the current dual-capsule ingestion protocol in this study. DNA extraction was performed on all 38/38 baseline SIMBA samples (1 baseline SIMBA and 1 post-intervention SIMBA were lost in feces), 28/40 post-intervention SIMBA samples and all fecal samples. 66 samples, including at least one from each ingestion time point, were allocated for 16S sequencing and a remaining 12 SIMBA samples were stored at -80°C for future analysis. 65/66 SIMBA capsules allocated for 16S sequencing had sufficient DNA of suitable quality for downstream analyses. In the 100 µL of the DNA elution, the median DNA concentration of baseline small intestine sample collected from SIMBA capsules was 0.058 ng/µL (IQR 0.039 – 0.082 ng/µL). The median DNA amount of post-intervention small intestine sample collected from SIMBA capsules was 0.557 ng/µl (IQR 0.153 – 1.41 ng/µl), which represents the high concentration of probiotics in the capsules. In contrast, the fecal samples contain much more concentrated DNA: median 561.13 ng/µL (IQR 491.28-688.34 ng/µL) for the baseline fecal DNA concentration and 629.20 ng/µL (IQR 519.38- 698.11 ng/µL) for post-intervention fecal DNA concentration.

### Microbiota composition is different between SIMBA and stool samples

Samples were analysed using a PCoA with an unsupervised Weighted UniFrac algorithm revealing 3 main clusters based on spatial disposition (Figure 4A). The orange (Baseline) and green (Probiotics) samples are concentrated on the left-center of the PCoA space and correspond to stool samples. The capsule samples are located on the right side and separated along the PC2 axis, suggesting group differences between stool samples and capsules, and between Capsules-Probiotics (red, bottom-right) and the Capsules-Baseline (blue, top-right). These differences are also visible on the grouped taxonomic bar plots (Figure 4B), where the apparent difference between Capsules-Baseline and Capsules-Probiotics is caused by the high number of *Lactobacilli* and *Bifidobacteria* from the probiotic co-ingestion. Indeed, the removal of *Lactobacilli* and *Bifidobacteria* from the grouped bar plot analysis by bioinformatic filtering increased the similarity between the Capsules-Probiotics and the Capsules-Baseline in a manner similar to stool samples (Figure S1).

**Figure 4:**
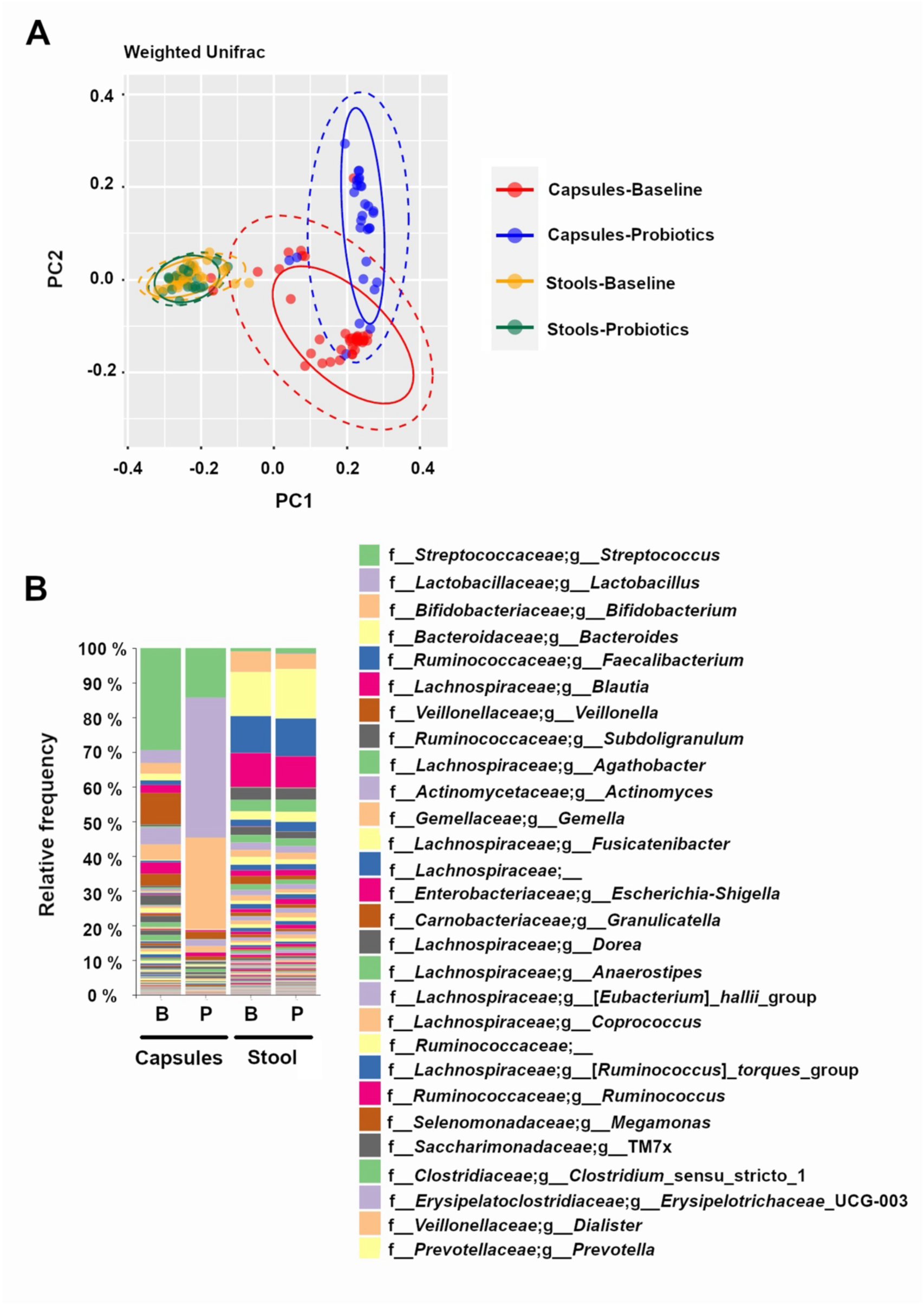
Differences between experimental groups shown on (A) a principal coordinate analysis (PCoA) with a weighted UniFrac algorithm (Orange = Stool- Baseline; Green = stool-Probiotics; Red = Capsule – Baseline; Blue = Capsule – Probiotics), and (B) on a grouped taxonomic bar plot showing genus-level relative abundances at baseline and with probiotics, for capsules and stool samples.

As expected for samples originating from the small intestine, alpha diversity measures were lower in the capsules compared to the stool samples (Figure S2), while the Capsules-Probiotics showed an even lower diversity than the Capsules-Baseline that is most likely due to the presence of high amounts of probiotic *Lactobacilli* and *Bifidobacteria*.

### Comparisons between groups by machine learning confirms the difference between SIMBA and stool samples

The overall differences between the stool and capsule samples were also assessed by machine learning group comparisons (Figure 5A) with 3 comparisons. The first comparison (Comp 1) included all the stool *vs*. capsules samples without consideration for the probiotic intervention. The very high final accuracy at 100% means that from training on genus-level taxonomic tables, the algorithm was always able to distinguish between the stool and capsules samples (i.e. the group differences were clear and the capsules are consistently different from the stool samples) (Figure 5B). In comparison 2 (Comp 2), a final accuracy of 45% (accuracy ratio of 0.83) means that the algorithm could not distinguish between stool-baseline or stool-probiotics very efficiently (Figure 5C). In comparison 3 (Comp 3), a final accuracy of 100 % shows that the algorithm was able to clearly distinguish between the capsule-baseline and capsule-probiotics (Figure 5D).

**Figure 5:**
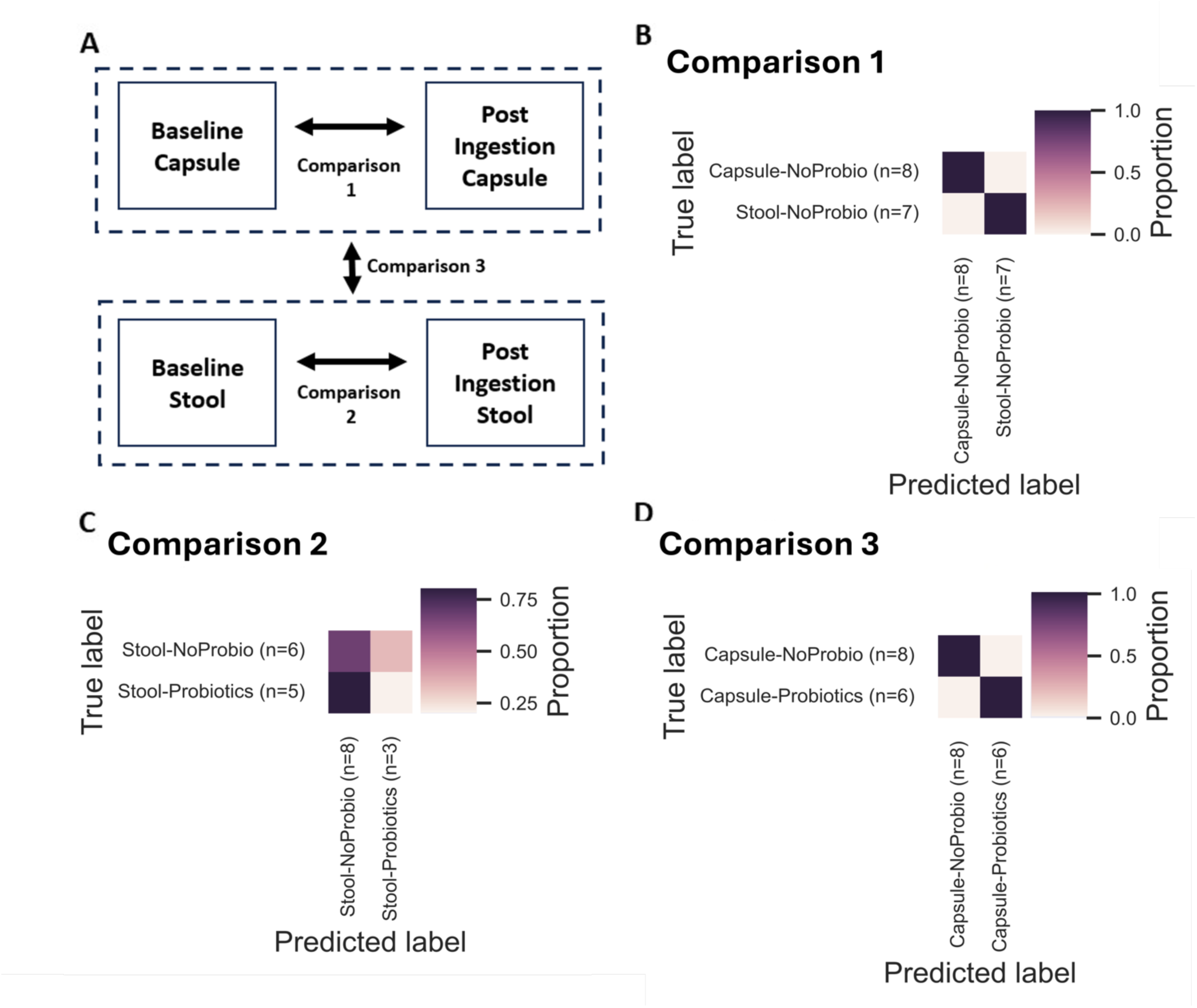
Summary of the pairwise group comparisons using Machine Learning Classifiers (as a QIIME2 module) using the ExtraTreesAlgorithm. For each of the 3 comparisons illustrated in (A), the confusion matrices are presented in (B) for Comparison 1 (Comp1; All samples – Stool *vs* Capsules; Final accuracy 100%, Accuracy ratio 1.79), in (C) for Comparison 2 (Comp2; Stool Baseline *vs* Stool Probiotics; Final accuracy 45%, Accuracy ratio 0.83), and in (D) for Comparison 3 (Comp3; Capsule Baseline *vs* Capsule Probiotics; Final accuracy 100%, Accuracy ratio 1.7).

### The main group classifiers are representative of their sampling region

Among the main classifiers identified by the machine learning algorithm (comparing Baseline stool samples versus Baseline SIMBA), several taxa previously associated with the SI microbiome were identified as enriched in the SIMBA capsules while taxa known to be associated with the colon were enriched in the stool samples (Figure 6). For example, *Streptococcus*, *Veillonella*, *Actinomyces*, *Gemella* and TM7x were enriched in the SIMBA capsule baseline sample, while *Bacteroides*, *Blautia*, *Faecalibacterium*, *Dorea* and *Anaerostipes* were enriched in the stool baseline samples.

**Figure 6:**
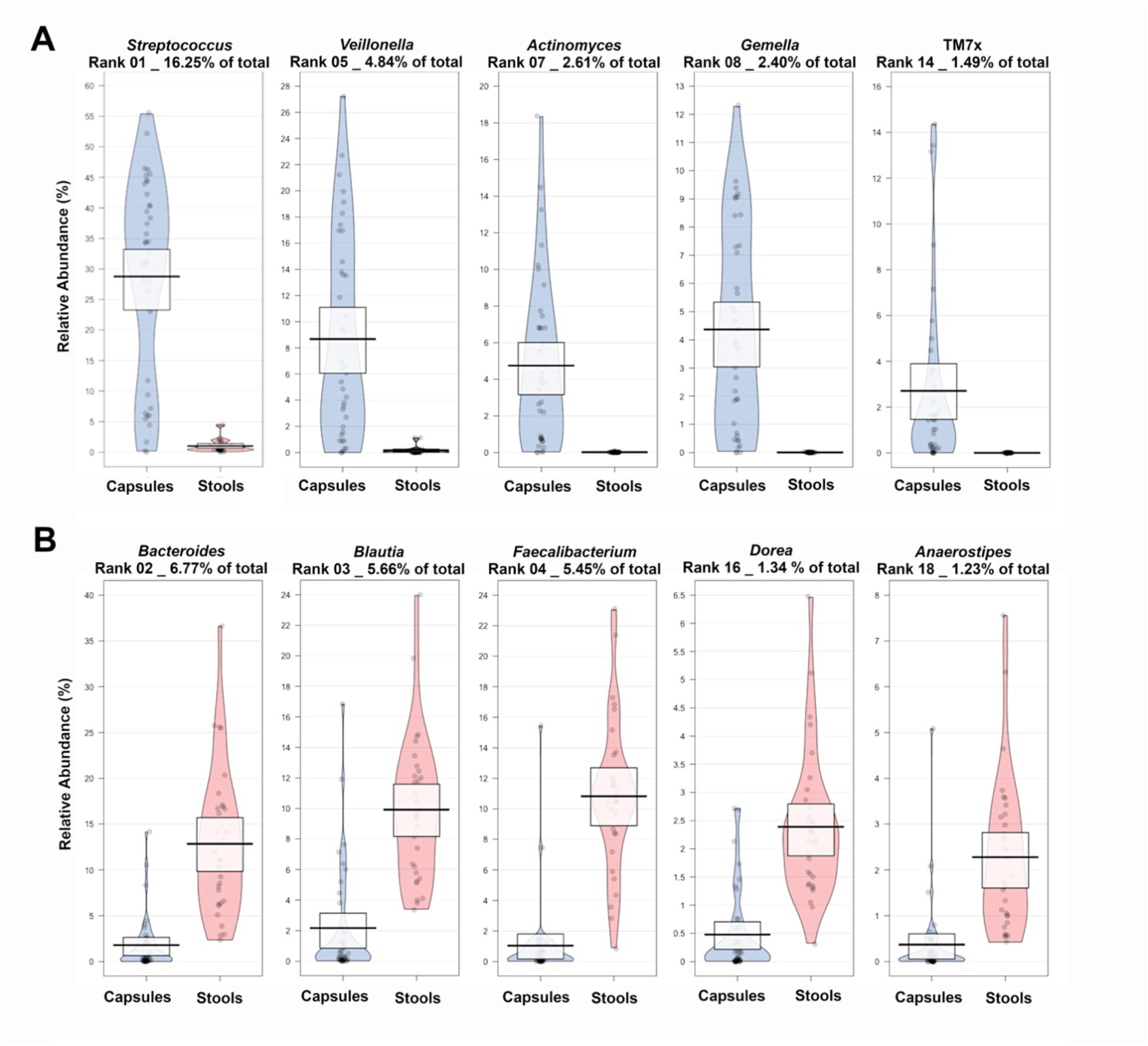
Pirate plots showing a selection of most abundant taxa enriched in (A) the capsules or in (B) the stool samples at baseline for the comparison between Baseline Capsule *vs* Baseline stool samples.

## Discussion

This study introduces the Small Intestine MicroBiome Aspiration (SIMBA) capsule, a minimally invasive device for collecting small intestine luminal fluid for microbiome analysis. SIMBA will allow to address a critical research gap in gastrointestinal microbiome studies, where the upper gastrointestinal tract’s unique characteristics are often overlooked due to the challenges of accessing and sampling this region.

*In vitro*, the capsules displayed an excellent sealing efficacy under conditions mimicking those encountered during a clinical trial (i.e., gut transit at 37 °C in fecal slurry and storage/shipping at 4 °C or -20 °C). For the preserving agent efficacy, our *in vitro* testing was very challenging, designed as a worst-case scenario with 6 days at 37 °C after manipulations for manual inoculations that are an unusual capsule usage. We observed that only 5 capsules out of 30 displayed a variable amount of staphylococcus contamination. However, it was clear with the negative controls that the preserving agent was successful at maintaining the community architecture of most samples in the absence of an external contamination. In a real-life setting, the capsule parts inside of their pH-sensitive outer shell are sterile and undergo DNA removal process during the manufacturing process. In the current experimental setting, it is likely that the capsules were contaminated with various amounts of *Staphylococci* during the inoculation or extraction of the cargo by pipetting, before the 16S amplification.

In the clinical study, the SIMBA capsule showed a remarkable ability to collect samples from the small intestine using the multiple X-ray tracking method, 97.2% (35/36) completing sample collection in the targeted region. The companion X-ray tracking method provides a relatively affordable and minimally invasive approach to validate the timing and location of sampling with our passive sampling capsule technology in our groundbreaking efforts to expand our understanding of the intestinal microbiome. Gradually, we aim to further enhance the accuracy of our capsule-based sampling technology and also to develop other means of capsule tracking for deploying the capsule to large-scale studies.

Microbiome analysis revealed significant differences between the small intestine microbiome profiles obtained with SIMBA and fecal microbiome profiles. This highlights the importance of directly sampling the small intestine for a more accurate understanding of diet-microbiota interactions. This technology possesses the potential to transform our comprehension of the gut microbiome and its implications in health and disease. Furthermore, it facilitates interventional studies for monitoring both the immediate and prolonged impacts on the small intestine microbiome resulting from various medications and nutraceutical products, including prebiotics, probiotics, and postbiotics. The capsule captured the co-ingested intervention, here a probiotic, which took a lot of space in the SIMBA capsule and appeared clearly in the bar plots of the microbiome analysis. This is interesting because probiotic interventions are reputably difficult to monitor after one dose using stool samples; this study proved that the probiotic bacteria reached the small intestine. In future probiotics studies, care should be given to allow enough time for washout of the probiotic bolus before ingesting SIMBA in order to study the effects of repeated doses on the SI microbiome. We have succeeded in removing the probiotic signal by filtering out all the *Bifidobacteria* and *Lactobacilli*, but this is an artificial analysis shown here as a proof-of-concept and the validity of using this approach to further analyze microbiome composition after filtering the samples should be determined.

When analyzing the baseline SIMBA and stool samples, there was a clear difference in microbiome composition between the 2 sampling locations, and the main genera enriched in the SIMBA capsule converge with the published microbiome composition of small intestine endoscopy samples. (21) There is still a lack of direct comparison with small intestine samples collected by other means, which is a limitation of this study. As the next step, samples collected from our SIMBA capsules will be compared against endoscopic aspirate samples.

The SIMBA capsules may offer a minimally invasive alternative to endoscopic aspiration, making it more comfortable for study participants while providing comprehensive spatial representation along the gastrointestinal tract. Its embedded preservative agent ensures the retention of time-stamped microbiome snapshots, crucial for studying dynamic effect of a concurrent intervention to the small intestine microbiome, either pharmaceutical or dietary.

## Conclusions

By addressing the need for minimally invasive devices to collect, seal and preserve the small intestine sample, the SIMBA capsule opens new avenues for research into various digestive conditions, such as small intestine bacterial overgrowth, irritable bowel syndrome, obesity, metabolic diseases, and cancer. This technology has the potential to transform our understanding of the gut microbiome and its role in health and disease.

This study’s results suggest that the SIMBA capsule is ready for use in studies aiming to elucidate microbiome-diet interactions. Further investigations involving diseased populations are ongoing to compare the results with fresh small intestine samples collected by endoscopic aspirates. In conclusion, the SIMBA capsule represents a significant advancement in microbiome research, complementing traditional fecal sampling procedures and expanding our capabilities to explore the complexities of the gastrointestinal microbiome.

## Data Availability

The datasets presented in this article were uploaded on the Sequence Read Archive (SRA) under the accession number BioProject ID: PRJNA1031660.

https://www.ncbi.nlm.nih.gov/bioproject/?term=PRJNA1031660

## Author Contributions

G.W., S.Br., T.A.T., S.Bi, C.N.A, conceived and planned the experiments. G.W. S.M. L.W. R.R. L.L., C.T., J.A., and O.M. carried out the experiments. S.M. planned and carried out the in-vitro simulations. S.M. and O.M. contributed to sample preparation. G.W., S.Br., L.L., S. Bi, J.A., T.A.T. and C.N.A. contributed to the interpretation of the results. G.W. took the lead in writing the manuscript. All authors provided critical feedback and helped shape the research, analysis and manuscript.

## Supplementary Material

Figures S1, S2 and S3.

**Supplementary Figure 1.**
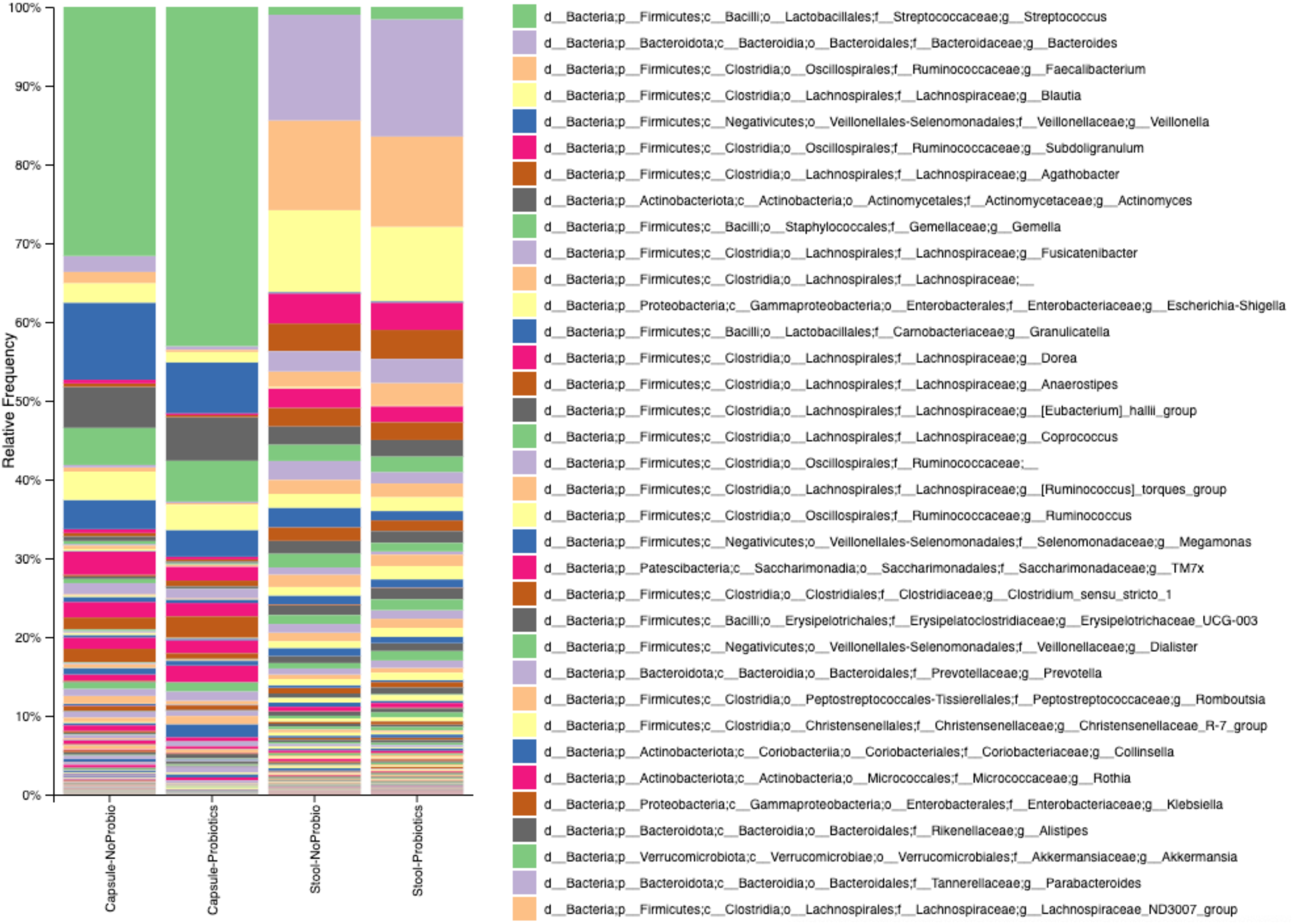
Grouped taxonomic bar plots showing genus level microbiome composition after removal of the *Lactobacilli* and *Bifidobacteria* from all groups.

**Supplementary Figure 2.**
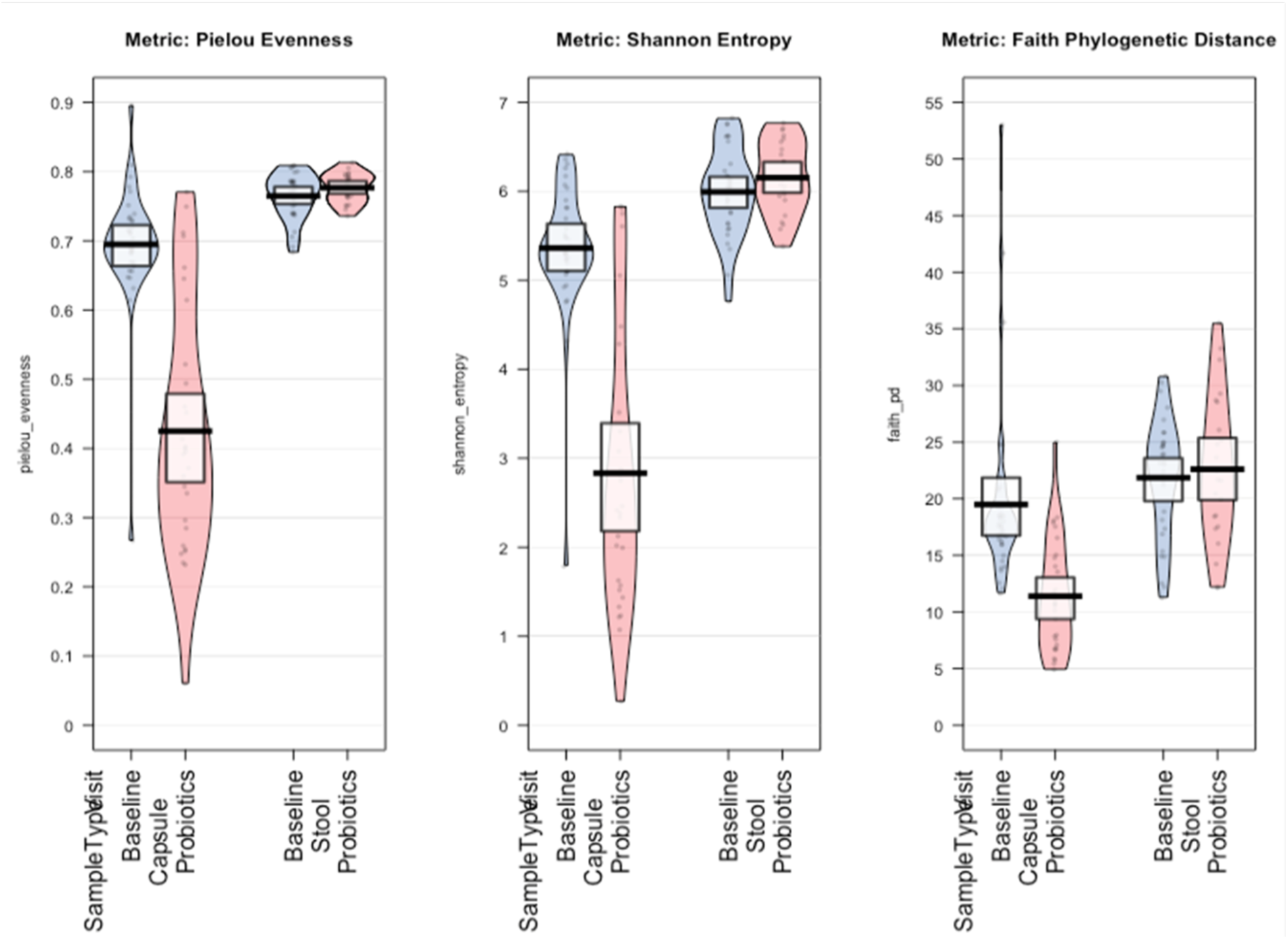
Alpha diversity metrics.

**Supplementary Figure 3.**
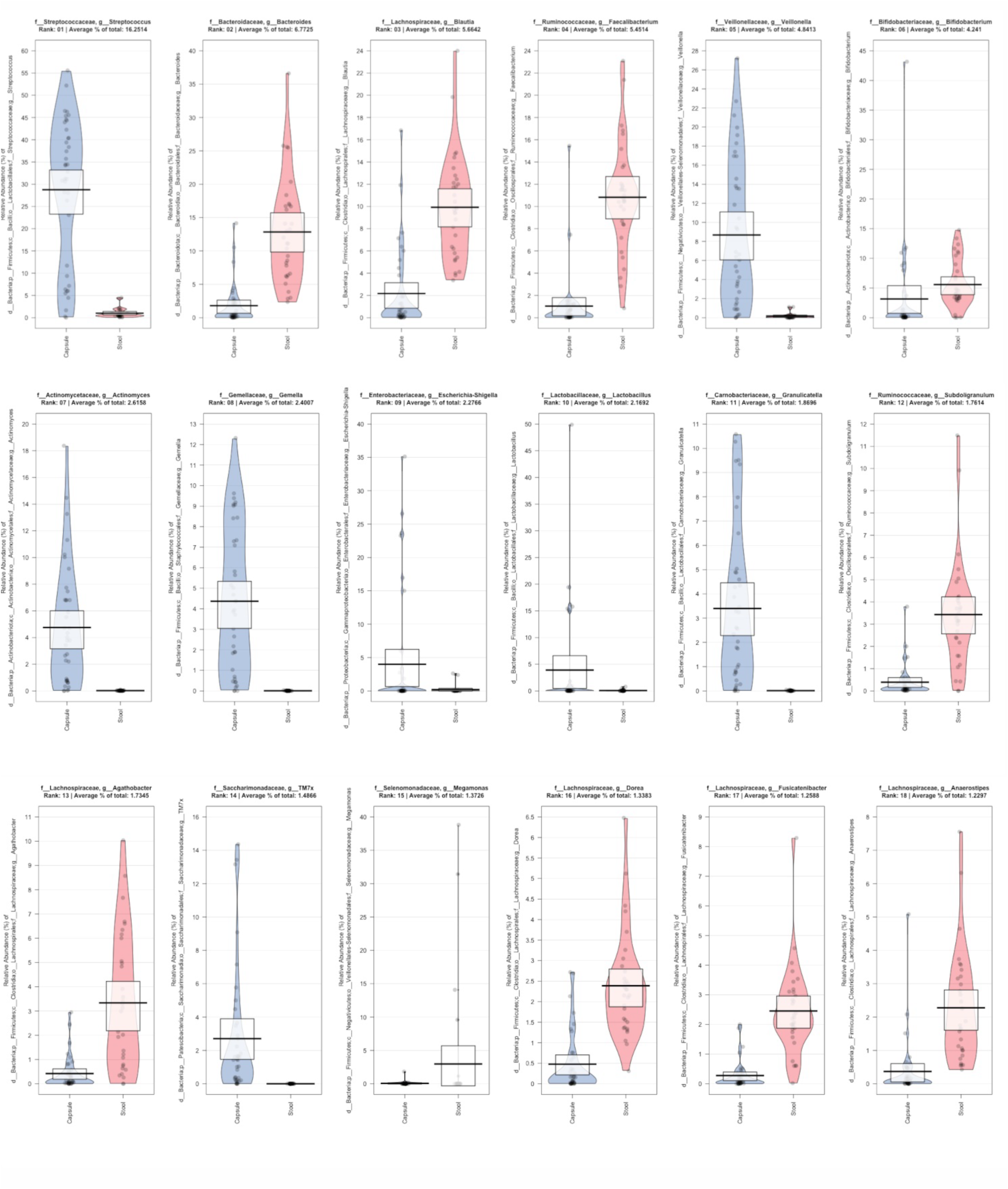
Top 18 classifiers of the comparison between baseline capsule and stool samples.

